# Influence of socioeconomic factors on cancer incidence and stage of melanoma in China

**DOI:** 10.1101/2019.12.19.19013706

**Authors:** Leqi Qian, Jiaqi Liu

## Abstract

**Background:** A high socioeconomic status (SES) was considered as an established risk factor for melanoma in western countries and areas, the same was not true in China. There have been few studies on SES of Chinese melanoma patients. The purpose of this study was to evaluate the association of SES in Chinese melanoma patients.

**Methods:** We performed a cross-sectional study using the data from Ministry of Human Resources and Social Security of the People’s Republic of China, and National Healthcare Security Administration. The clinical stage at diagnosis was categorized according to Guidelines of Chinese Society Oncology, Melanoma.

**Results:** We enrolled 122 patients with pathologic melanoma in Zhongshan Hospital, Fudan University between January 2013 to December 2017. 58 (48%) were male and 64 (52%) were female, the mean age was 59.23±9.91 years and median age was 60.5 years. Patients in 45-59 age group and 60-73 age group tend to have higher cancer incidence compared with other age groups. The acral lentiginous melanoma (48%) was the most common subtype. Patients with low education level (middle school and below) and low monthly household income (< 3000CNY) were associated with a greater risk of melanoma. Similar results were indicated for the patients who were unemployed. There were remarkable more patients who had medical insurance diagnosed with melanoma in this study. There was no significant difference on melanoma stage for patients with higher compared to lower education level (*p*=0.153). For monthly household income, the Fish’s exact test indicated no significant difference in melanoma stage with *p*=0.507. No staging difference was observed between unemployed patients and employed patients (*p*=0.687). Finally, statistically significant differences in melanoma staging were not indicated by a patient’s insurance status (*p*=0.537).

**Conclusions:** For patients enrolled in our study, disadvantaged SES did not substantially lead to an increasing risk of melanoma incidence, and the effect of socioeconomic factors seemed uninfluenced the stage of melanoma.

---

Melanoma rates have risen dramatically all over the world in the last 15 years, with steady increases seen in cases of invasive melanoma with higher morbidity and mortality rates(1).

In China Melanoma represents a small proportion of tumors, it is nevertheless one of the main cancers with high mortality(2). Melanoma has also been one of the most rapidly increasing cancers in China(3).

According to reports based on western data, melanoma has consistently been shown to occur frequently in groups of high socioeconomic status (SES)(4-7). For example, analysis of the United States data demonstrated higher incidence rates of melanoma in counties with high richness rates, high education level, high median household income and high employment rates(8). This association has mainly been attributed to high-risk behavioral patterns of excessive sun exposure, as patients with higher income and education levels have more opportunity for recreational sunlight(9, 10). However, in China, the most common pathological subtype of melanoma, differed substantially from those reported in western populations, is the acral lentiginous which does not appear to be related to sun exposure(11). Meanwhile, the rarity of melanoma in China precludes in-depth and intensive economic and social studies of this disease. There have been few studies on SES statues of melanoma patients in China. Thus, to date, the impact of SES among Chinese melanoma patients is not well defined. To bridge the gaps of extant literatures, this study aimed to retrospectively analysis the SES of melanoma patients documented in our institution, to explore the association between SES and melanoma in China through a representative sample.

## Patients and Methods

### Patients

This study was conducted at Zhongshan Hospital, Fudan University, which is a tertiary hospital and a budget management unit under the China National Health and Family Planning Commission. This hospital has a wide national coverage, thus its patients we included could be regarded as a good sample representing a national situation. This study adhered to the guidelines set of the declaration of Helsinki, and was approved by the institutional review board of Zhongshan Hospital. All patients provide informed written consents.

We studied a cohort of patients who presented to Zhongshan hospital with diagnosis of melanoma, first and primary, over a period of 5 years between January 2013 to December 2017. Patients with unclear medical records and incomplete socioeconomic information were excluded. Patients who had undergone diagnosis or therapy in other facility centers were also excluded due to the difficulties in retrieving information.

Patients’ demographics including ID number (which contains the date of birth), gender and age were obtained at time of enrollment. Age was categorized into five groups: younger than 30 years, 30-44 years, 45-59 years, 60-74 years, and 75 years or older according to a national wide research of China(12).

SES were estimated using data from Ministry of Human Resources and Social Security of the People’s Republic of China (MOHRSS) and National Healthcare Security Administration (NHSA). Sociodemographic data included education, ethnicity, and phone number. Economic data included household income per capita, occupation, and medical insurance. Date of pathologic diagnosis was defined as the biopsy date of the primary tumor and was used to calculate each patient’s age at melanoma diagnosis.

Patients’ tumor characteristics were obtained from hospital information system (HIS) of Zhongshan hospital. The histological subtypes and the clinical stage at diagnosis were categorized according to Guidelines of Chinese Society Oncology (CSCO), Melanoma (2017. V1). Tumors were grouped into five histological subtypes: superficial spreading malignant melanoma (SSM), lentigo maligna melanoma (LMM), nodular melanoma (NM), acral lentiginous melanoma (ALM) and others (mucosal, unclassified and unknow).

### SES variables

Variables of SES for analyses included social and economic characteristics as recorded at time of diagnosis. We categorized SES variables according to previous Chinese researches. The educational status was categorized into two groups: middle school and below; high school and above(13). Monthly household income was categorized into two levels: <3000 CNY (approximate 430 USD); ≥3000 CNY(14). In this cohort, there was no patient whose occupation was professor or semi-professor, thus occupational status was categorized into two levels: unemployed and employed(15). Medical insurance status was coded as insured or uninsured. The Han ethnic group is the dominant one in China, accounting for approximate 90% over the whole nation population. Therefore, the ethnicity was binarily categorized Han and other groups. We defined that combination with high social and high economic status as high SES.

### Statistical Methods

Statistical analyses were conducted using SPSS software (version 22.0, IBM Institute, California, USA). Continuous variables were presented as mean ± standard deviation (SD) or median with range depending on their distribution. Categorical variables were presented as frequency and percent. Occurrence of stages melanoma was cross-classified separately by each categorical variable and was examined with Fisher exact test when appropriate. A *P*-value < 0.05 was considered statistically significant unless otherwise specified.

Interactions between single socioeconomic variables with sex and age were tested one pair at a time with Chi square statistics. No significant interaction existed.

## Results

Based on these inclusion and exclusion criteria, we identified a cohort of 122 patients with pathologic melanoma diagnosed.

Table 1 provides a summary of the demographic, socioeconomic and major tumor characteristics of the cohort. Of the 122 patients included, 58 (48%) were male and 64 (52%) were female, the mean age was 59.23±9.91 years and median age was 60.5 years. Patients in 45-59 age group and 60-73 age group tended to have higher cancer incidence compared with other age groups. In this cohort, there were remarkable more patients (46%) diagnosed with stage II cancer, vs stage III (28%), stage I (25%) and stage IV (2%). We found that the ALM (48%) was the most common subtype, followed by SSM (16%) and other subtypes (16%). NM (12%) and LMM (7%) histological types were relatively rare compared to ALM. Patients with low education level (middle school and below) and low monthly household income (<3000CNY) were associated with a greater risk of melanoma. Similar results were indicated for the patients who were unemployed. There were remarkable more patients who had medical insurance diagnosed with melanoma in our cohort.

**Table 1.** Patient demographic, socioeconomic and major tumor characteristics

|  | N | % |
| --- | --- | --- |
| Sex |  |  |
| Male | 58 | 48 |
| Female | 64 | 52 |
| Age of diagnosis (years) |  |  |
| ≤30 | 3 | 2 |
| 31-44 | 1 | 1 |
| 45-59 | 55 | 45 |
| 60-73 | 60 | 49 |
| ≥74 | 5 | 4 |
| CSCO stage |  |  |
| I | 31 | 25 |
| II | 56 | 46 |
| III | 34 | 28 |
| IV | 3 | 2 |
| Histological type |  |  |
| SSM | 20 | 16 |
| LMM | 9 | 7 |
| NM | 15 | 12 |
| ALM | 58 | 48 |
| Other | 20 | 16 |
| Educational level |  |  |
| Middle school and below | 74 | 61 |
| High school and above | 48 | 39 |
| Household income (monthly) |  |  |
| <3000 CNY | 87 | 71 |
| ≥3000 CNY | 35 | 29 |
| Occupational status |  |  |
| Unemployed | 69 | 57 |
| Employed | 53 | 43 |
| Medical insurance |  |  |
| Uninsured | 46 | 38 |
| Insured | 76 | 62 |
| Ethnicity |  |  |
| Han | 117 | 96 |
| Others | 5 | 4 |

The descriptive statistics are presented in table 2. The Fish’s exact test analyzed the association between the SES factors and melanoma stage. There was no significant difference on melanoma stage for patients with higher compared to lower education level (*p*=0.153). For monthly household income, the Fish’s exact test indicated no significant difference in melanoma stage with *p*=0.507. No staging difference was observed between unemployed patients and employed patients (*p*=0.687). Finally, statistically significant differences in melanoma staging were not indicated by a patient’s insurance status (*p*=0.537). Overall results indicated that no stage difference was observed between high SES and low SES.

**Table 2.**
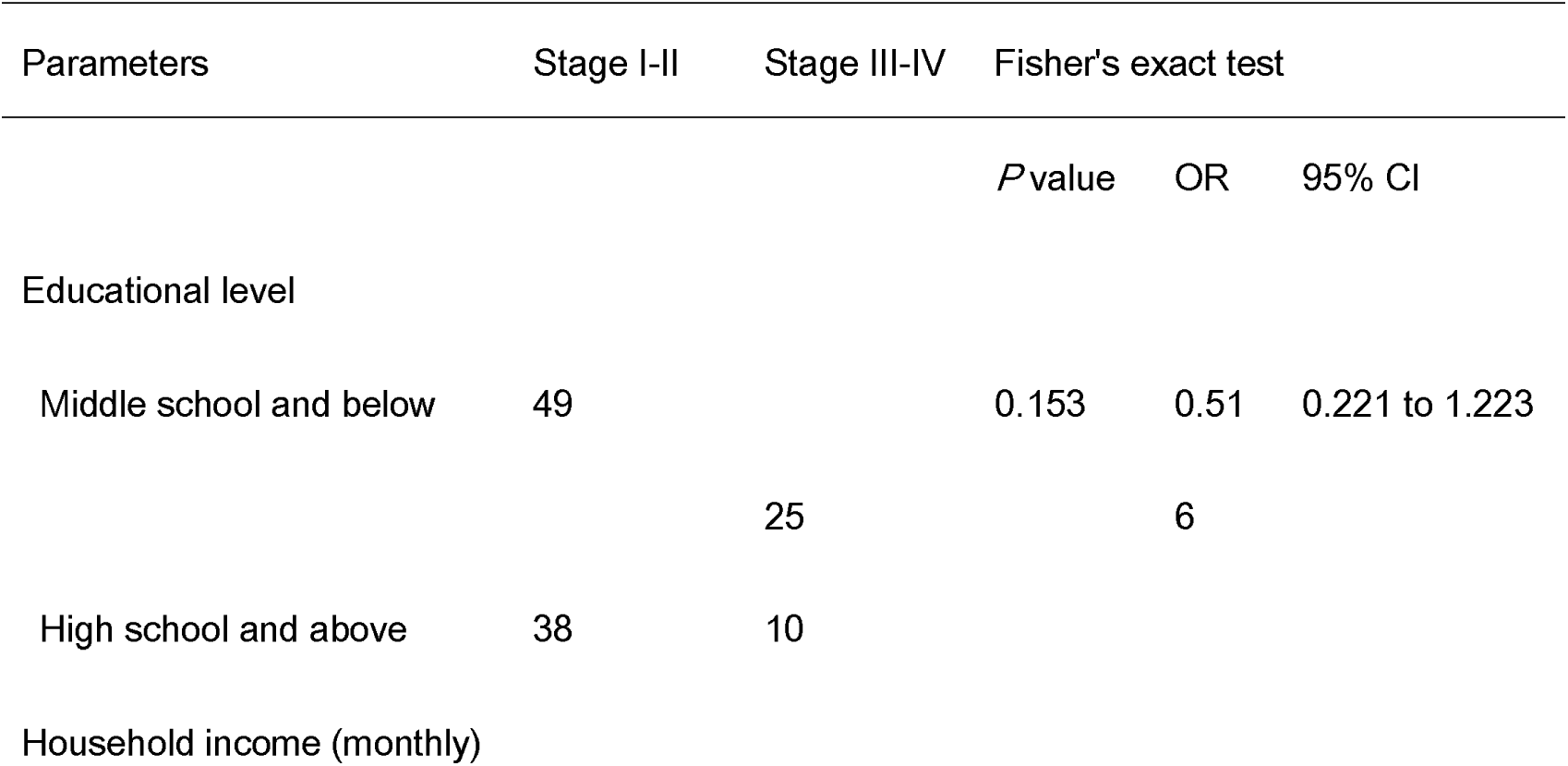
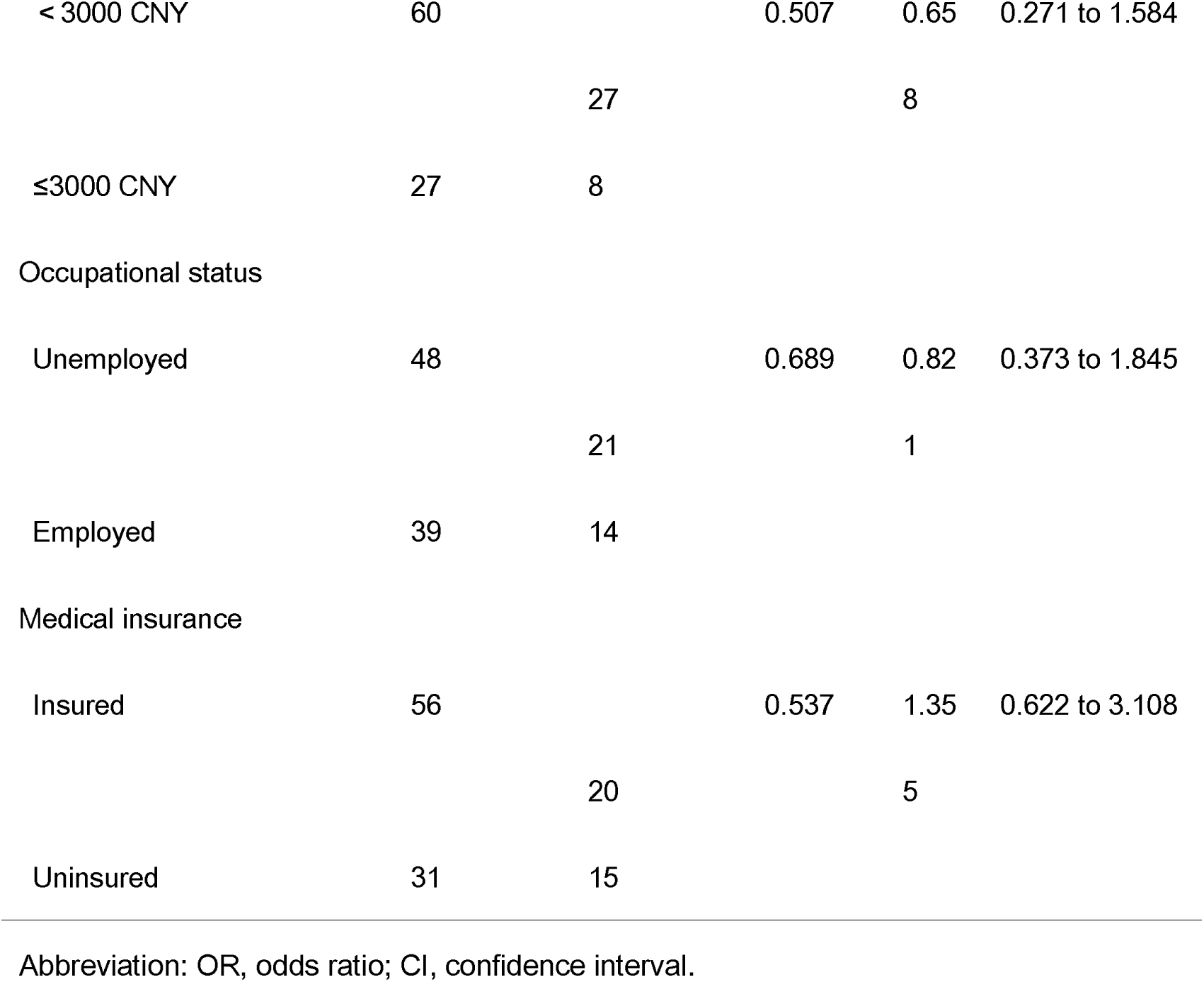
Associations between SES factors and melanoma stage.

| Parameters | Stage I-II | Stage III-IV | Fisher's exact test |  |  |
| --- | --- | --- | --- | --- | --- |
|  |  |  | *P*value | OR | 95% CI |
| Educational level |  |  |  |  |  |
| Middle school and below | 49 |  | 0.153 | 0.51 | 0.221 to 1.223 |
|  |  | 25 |  | 6 |  |
| High school and above | 38 | 10 |  |  |  |
| Household income (monthly) |  |  |  |  |  |
| < 3000 CNY | 60 |  | 0.507 | 0.65 | 0.271 to 1.584 |
|  |  | 27 |  | 8 |  |
| ≤3000 CNY | 27 | 8 |  |  |  |
| Occupational status |  |  |  |  |  |
| Unemployed | 48 |  | 0.689 | 0.82 | 0.373 to 1.845 |
|  |  | 21 |  | 1 |  |
| Employed | 39 | 14 |  |  |  |
| Medical insurance |  |  |  |  |  |
| Insured | 56 |  | 0.537 | 1.35 | 0.622 to 3.108 |
|  |  | 20 |  | 5 |  |
| Uninsured | 31 | 15 |  |  |  |

Abbreviation: OR, odds ratio; CI, confidence interval.

## Discussion

In general, melanoma has been shown to occur more frequently in Caucasian populations with an incidence rate of 29.4 per 100,000(1). In contrast, the incidence rates in China was 0.8 per 100,000(12), which represents a decreased incidence of at least 30 times when compared to non-Hispanic Whites. Multiple factors have been attributed to melanoma incidence, including environmental, social, and socioeconomic factors. Based on western researches, melanoma represented an area of interplay amongst ethnicity, socioeconomic factors, and incidence of disease. Specifically, melanoma has consistently been shown to occur more frequently in groups of higher SES(4, 16). However, we did not observe the evidence of association between SES factors and melanoma in China. Thus, this study is devoted to research the influence of socioeconomic factors on incidence and stage of melanoma in China

SES is a multidimensional concept and most contemporary researchers agree that it is represented by a combination of family income, education and occupational status(17). In this study, SES variables for analyses included educational level, monthly household income, occupational status and medical insurance status.

The SES data in this study was obtained from MOHRSS and NHSA. MOHESS coordinates human resources management, employment and social security policies. And MHSA sets up the budget of national medical insurance, and are responsible for the settlement of accounts. Trough checking patient ID number, we can obtain the information of insurance, education, income and occupation.

Generally speaking, regardless of the cancer subtypes, the best results come from early detection(18-20). In melanoma, early detection results in reduced morbidity and mortality(21). We can draw a conclusion that the likelihood of patients to actively self-detection is dependent on the patient’s education level, patients who are unable to perceive the risk of melanoma have increased difficulty in performing risk reduction actions. But this conclusion is still controversial. Pollitt *et al*, who linked lower education level with patients’ decreased perception of risk for melanoma, and they found that even physicians were less likely to inform patients with lower education level that they were at risk of skin cancer (22). Ibfelt *et al* also showed that patients who were socially disadvantaged in terms of education status had an increased risk of a diagnosis with advanced-staged melanoma(23). However, an United States study demonstrated higher incidence rates of melanoma in counties may associated with higher richness rates, higher education level, higher median household income and higher employment rates(8). This result could be explained by the fact that patients with higher education levels have more opportunity for sunlight(10). Our study indicated that lower education level was associated with a higher risk of melanoma incidence, but could not influence the melanoma staging. We believe that our result will be an important finding in melanoma research because, regardless the education level, Chinese always appreciate fair skin, we like to get away from sunlight so that we will not get tanned. This may explain the association between education level and melanoma epidemiology in our research.

In this study, the lower income status, unlike western results[18, 19], did not associate with the advanced stage of melanoma. The western results may be explained by the fact that patients with lower income could be affected by lack of resources, delayed detection, poorer quality of care, and fewer treatment options. In China, we did not have the clinical advance of melanoma treatment till 2018 when PD-1 blockage was approved by the China Food and Drug Administration (CFDA)(24). Before that the burden of melanoma treatment was not loaded due to the lack of targeted and immuno treatments. This limited treatment option led to a poor prognosis, meanwhile, minimized the effect of income. Patients did not need to pay too much for treatment.

While unemployment has been shown to be associated with a decreased relative risk of melanoma in western research(25), our study found that unemployed patients had higher melanoma incidence compared to employed but did not had influence on melanoma staging. In MOHRSS and NHSA, farming and freelancing may be classified as “unemployment”, we are not clear about the impact of this occupation classification on epidemiological outcomes.

Overall, unlike western results, analysis of our data demonstrated that lower SES status was not associated with an increased incidence in a statistically significant amount for all stages of melanoma.

In a rapid developing country, a longer period would inevitably be accompanied rapid advances in economic and society, this heterogeneity may lead to bias. Therefore, we retrospectively analyzed patients present to our hospital and diagnosed as melanoma between January 2013 and December 2017. In this 5-year period, we can establish a relatively large sample and minimize the socioeconomic disparities due to time variations. This study is not without limitations. A limitation in our result is that the MOHRSS and MHSA do not collect medical history and cancer specific survival, thus we did not explore the associate between SES and survival. Additionally, whereas this study enrolled 122 patients, a relatively large cohort in China, it was not population based. Further national wide studies should investigate.

## Conclusion

The results of the present study partially support our hypotheses, suggesting that there was no significant association among high SES, melanoma incidence and stage. Patients who were socially disadvantaged in terms of education, income, occupation or insurance status did not have an increased risk of melanoma incidence. The effect of socioeconomic factors seemed uninfluenced the cancer stage of melanoma in this study.

## Data Availability

All data, models, and code generated or used during the study appear in the submitted article.

## Notes

### Competing Interest Statement

The authors have declared no competing interest.

### Funding Statement

JL is currently receiving the grant (#81802724) from the National Natural Science Foundation of China. For the remaining authors none were declared.

## References

1. Siegel RL, Miller KD, Jemal A. Cancer statistics, 2018. CA Cancer J Clin 2018;68:7–30

2. Zhang M, Zhang N. Clinical and prognostic factors in 98 patients with malignant melanoma in China. J INT MED RES 2017;45:1369–1377

3. Si L, Zhang X, Shu Y, et al. A Phase Ib Study of Pembrolizumab as Second-Line Therapy for Chinese Patients With Advanced or Metastatic Melanoma (KEYNOTE-151). TRANSL ONCOL 2019;12:828–835

4. Harvey VM, Patel H, Sandhu S, et al. Social determinants of racial and ethnic disparities in cutaneous melanoma outcomes. CANCER CONTROL 2014;21:343–349

5. Jiang AJ, Rambhatla PV, Eide MJ. Socioeconomic and lifestyle factors and melanoma: a systematic review. Br J Dermatol 2015;172:885–915

6. Schadendorf D, van Akkooi A, Berking C, et al. Melanoma. LANCET 2018;392:971–984

7. Broadbent T, Bingham B, Mawn LA. Socioeconomic and Ethnic Disparities in Periocular Cutaneous Malignancies. SEMIN OPHTHALMOL 2016;31:317–324

8. Singh SD, Ajani UA, Johnson CJ, et al. Association of cutaneous melanoma incidence with area-based socioeconomic indicators-United States, 2004-2006. J AM ACAD DERMATOL 2011;65:S58–S68

9. Chang C, Murzaku EC, Penn L, et al. More skin, more sun, more tan, more melanoma. AM J PUBLIC HEALTH 2014;104:e92–e99

10. Ortiz CA, Goodwin JS, Freeman JL. The effect of socioeconomic factors on incidence, stage at diagnosis and survival of cutaneous melanoma. Med Sci Monit 2005;11:A163–A172

11. Darmawan CC, Jo G, Montenegro SE, et al. Early Detection of Acral Melanoma: A Review of Clinical, Dermoscopic, Histopathologic, and Molecular Characteristics. J AM ACAD DERMATOL 2019

12. Chen W, Zheng R, Baade PD, et al. Cancer statistics in China, 2015. CA Cancer J Clin 2016;66:115–132

13. Chen C, Cheng G, Pan J. Socioeconomic status and breastfeeding in China: an analysis of data from a longitudinal nationwide household survey. BMC PEDIATR 2019;19

14. Xiao W, Ye H, Zeng H, et al. Associations among Socioeconomic Factors, Lag Time, and High-Risk Histopathologic Features in Eyes Primarily Enucleated for Retinoblastoma. CURR EYE RES 2019:1–6

15. Fuligni AJ, Zhang W. Attitudes toward family obligation among adolescents in contemporary urban and rural China. CHILD DEV 2004;75:180–192

16. Ortiz CAR, Goodwin JS, Freeman JL. The effect of socioeconomic factors on incidence, stage at diagnosis and survival of cutaneous melanoma. Medical science monitor : international medical journal of experimental and clinical research 2005;11:A163

17. Conger RD, Donnellan MB. An interactionist perspective on the socioeconomic context of human development. ANNU REV PSYCHOL 2007;58:175–199

18. Ozer ED, Suna N, Boyacioglu AS. Management of Hepatocellular Carcinoma: Prevention, Surveillance, Diagnosis, and Staging. EXP CLIN TRANSPLANT 2017;15:31–35

19. Barocas DA, Chang SS. Penile cancer: clinical presentation, diagnosis, and staging. Urol Clin North Am 2010;37:343–352

20. Deslauriers J, Pearson FG, Shamji FM. Recent advances in screening, diagnosis, and staging of lung cancer: a tribute to Robert J. Ginsberg. THORAC SURG CLIN 2013;23:xiii–xiv

21. Yagerman S, Marghoob A. Melanoma patient self-detection: a review of efficacy of the skin self-examination and patient-directed educational efforts. Expert Rev Anticancer Ther 2013;13:1423–1431

22. Pollitt RA, Swetter SM, Johnson TM, et al. Examining the pathways linking lower socioeconomic status and advanced melanoma. CANCER-AM CANCER SOC 2012;118:4004–4013

23. Ibfelt EH, Steding-Jessen M, Dalton SO, et al. Influence of socioeconomic factors and region of residence on cancer stage of malignant melanoma: a Danish nationwide population-based study. 2018;Volume 10:799–807

24. Li Z, Song W, Rubinstein M, et al. Recent updates in cancer immunotherapy: a comprehensive review and perspective of the 2018 China Cancer Immunotherapy Workshop in Beijing. J HEMATOL ONCOL 2018;11:142

25. Reyes-Ortiz CA, Goodwin JS, Freeman JL, et al. Socioeconomic status and survival in older patients with melanoma. J AM GERIATR SOC 2006;54:1758–1764

